# Burnout among healthcare professionals during COVID-19 pandemic: a cross-sectional study

**DOI:** 10.1101/2020.06.12.20129650

**Authors:** Mohammad Jalili, Mahtab Niroomand, Fahimeh Hadavand, Kataun Zeinali, Akbar Fotouhi

**Author notes:** Corresponding Author Address: 7th Floor, Bldg. No.2 SBUMS, Arabi Ave, Daneshjoo Blvd, Velenjak, Tehran, Iran., Postal Code: 19839-63113. **Financial Support:** None.

## Abstract

**Background:** The unpredictable nature of the new COVID-19 pandemic and the already alarming incidence of healthcare workers being affected can have a significant impact on the psychological well-being of the staff.

**Objective:** To describe the prevalence of burnout among healthcare professionals and the associated factors.

**Design:** Cross-sectional survey.

**Setting:** Eight university affiliated hospitals in the capital city of Tehran, Iran.

**Participants:** All healthcare workers at the study sites who had been taking care of COVID-19 patients.

**Measurements:** Age, gender, marital status, having children, hospital, job category, experience, and work load, as well as the level of burnout in each subscale.

**Results:** 326 persons (53.0%) experienced high levels of burnout. The average score in emotional exhaustion, depersonalization and lack of personal accomplishment was 26.6, 10.2, and 27.3, respectively. The level of burnout in the three subscales varied based on the personal as well as work related factors and gender was the only variable that was associated with high levels of all three domains.

**Limitations:** There was no control group and thus we cannot claim a causal relationship between COVID-19 and the observed level of burnout. Not all confounding factors might have been accounted for.

**Conclusions:** Burnout is prevalent among healthcare workers caring for COVID-19 patients. Age, gender, job category, and site of practice contribute to the level of burnout that the staff experience.

**Funding source:** None

## Introduction

Since December 2019, when the first cases of human infection with the novel coronavirus occurred (1), the emerging infectious disease (now called COVID-19) has rapidly spread worldwide, affecting people in 210 countries and territories with the current tally exceeding 7.5 million infected people and more than 423,000 deaths (2). In addition to the lives it has claimed globally, the pandemic has led to high levels of panic and anxiety throughout the world (3-6).

Although varying in different countries, healthcare workers comprise a notable proportion of the people who contracted the illness. According to some reports, 10% of confirmed cases of COVID-19 involved healthcare providers (7). A recent report by health authorities of Iran mentioned that around 10,000 healthcare professionals have contracted COVID-19 and at least 43 are known to have died of this disease (8). This high rate of infection and mortality has a tremendous impact on the healthcare system.

Burnout is a psychological syndrome first described by Maslach et al (9) and defined as a state of psychological, emotional, and physical stress in response to prolonged exposure to occupational stress. It includes feelings of emotional exhaustion (depletion of emotional resources), depersonalization (developing cynical attitudes about patients), and reduced professional accomplishment (a sense of negative evaluation of oneself).

The deadly and uncontrollable nature of COVID-19, for which there is no known effective treatment, together with relatively high rate of infection and mortality among healthcare providers can provoke the feelings of anxiety and stress in medical staff. Issues such as social stigmatization, shortage of personal protection equipment supplies, and heavy workload on the staff can aggravate this situation. Therefore, this pandemic is expected to have substantial psychological impact on healthcare providers (10). Burnout can have serious consequences for both patients and the healthcare professionals. It not only results in poor physical and mental health outcomes, lack of motivation, absenteeism, and low morale in the staff, but also leads to deterioration of the quality of care provided by the affected staff with resulting poor outcomes for patients. Several systematic reviews of the literature have found that high levels of burnout in health care professionals are associated with less-safe patient care (11,12). These consequences impose immense costs on the society (13,14).

Health authorities need more information on the magnitude of this problem and its associated factors in order to better prepare for future infectious disease outbreaks, and also to adapt sound interventions and strategies to prevent further deepening of this dreadful situation. Identification of risk factors which make certain professionals more susceptible to burnout would be an important step. These may include outbreak exposure and work related factors as well as personal characteristics.

This study was conducted on healthcare workers providing care for COVID-19 patients and aimed to explore the level of burnout among this population and also to examine factors associated with the development of this psychological sequel.

## Methods

### Study Design and Participants

This cross-sectional study was conducted during the current COVID-19 pandemic to evaluate the level of burnout among healthcare providers who were taking care of covid-19 patients at 8 university-affiliated hospitals. Altogether, these hospitals cover about one-third of Tehran province with a catchment area of about 4000000. The study was conducted about 2 months after the onset of the outbreak and when the disease had not yet been controlled.

Study participants included all healthcare providers (i.e. physicians, residents, interns, and nurses) who had taken care of COVID-19 patients at anytime during the first 2 months of the outbreak. Exclusion criteria were those with no patient contact during the outbreak or those unwilling to participate.

### IRB Approval

The Institutional Review Board granted approval and the requirement for written informed consent was waived based on the recognition that answering the survey instrument implied consent. Participation was voluntary and anonymity was assured. All personal information was kept confidential. Furthermore, researchers analyzed only de-identified data.

### Data Collection

The data collection instrument comprised of two parts: The first part of the tool asked questions pertaining socio-demographic and work-related characteristics. Participants were requested to indicate their age, gender, marital status, number of children, job title, place of work, and years of experience. This section also asked whether the respondent had been involved in the care of corona patients and if yes for how many shifts a month and how many hours per shift. The second part of the study tool was a translated version of Maslach Burnout Inventory (MBI) for Human Services Survey. In order to limit the study to burnout related to COVID-19, the phrase “due to COVID-19” was added to each item. MBI is an internationally recognized, validated, self-report questionnaire for measuring the severity of workplace burnout (15), using the three dimensions of emotional exhaustion, depersonalization, and personal accomplishment. The questionnaire has 22 items and each item is answered on a five-point Likert scale. This tool has been extensively used in many studies in different parts of the world and the Persian translation has also been validated previously (16).

The study used a convenience sampling method for recruitment. Invitation to participate in the study was made through professional as well as informal networks. The message included an invitation explaining the purpose of the study, the name and contact details of the principal investigator, and a live link to the host survey platform (E Poll). The online self-administered questionnaire instructed participants to respond to the statements in the tool in relation to the COVID-19 outbreak. Verbal reminders through face-to-face contact and phone calls were performed by a site coordinator at each hospital.

### Statistical Analysis

Burnout is expressed by scores of each of the three MBI subscales, with a high score meaning a high level of burnout. Each subscale score is calculated by adding up all scores of all items in that subscale, with the notion that the items on personal accomplishment domain are reversely scored (9,15). Scores range from 0 to 36 for emotional exhaustion, from 0 to 20 for depersonalization, and from 0 to 32 for personal accomplishment subscale. The standard cut-off values were used to define low, moderate, and high levels in each dimension (9,15).

We calculated the “average daily workload” by multiplying the number of shifts per month by the number of work hours per shift divided by 30 (number of days of a month). We also defined “high burnout” as obtaining a moderate or high score in either emotional exhaustion or depersonalization axes.

Upon completion of the study period, the data was downloaded from the online survey tool entered into an excel spreadsheet and anonymized. This data was, then, imported into the statistical software for analysis.

Descriptive analyses were conducted to examine all baseline characteristics of the participants and the outcome variables. Means and standard deviations (SDs) were calculated for continuous variables, while frequencies and percentages were produced for categorical variables. All reported p values are 2-tailed, and p < 0.05 was considered statistically significant. In order to explore the risk factors associated with the development of burnout, multivariable logistic regression models were used. Odds ratios and corresponding 95% confidence intervals (CI) were reported.

## Results

We invited a total of 1,002 healthcare workers to participate in the study, from which 645 people completed the survey (response rate: 64.4%). Thirty respondents declared that they had not taken care of COVID-19 patients, leaving a total of 615 completed questionnaires for analysis.

Table 1 illustrates the baseline characteristics as well as the mean MBI scores in each subscale in the studied population.

**Table 1.**
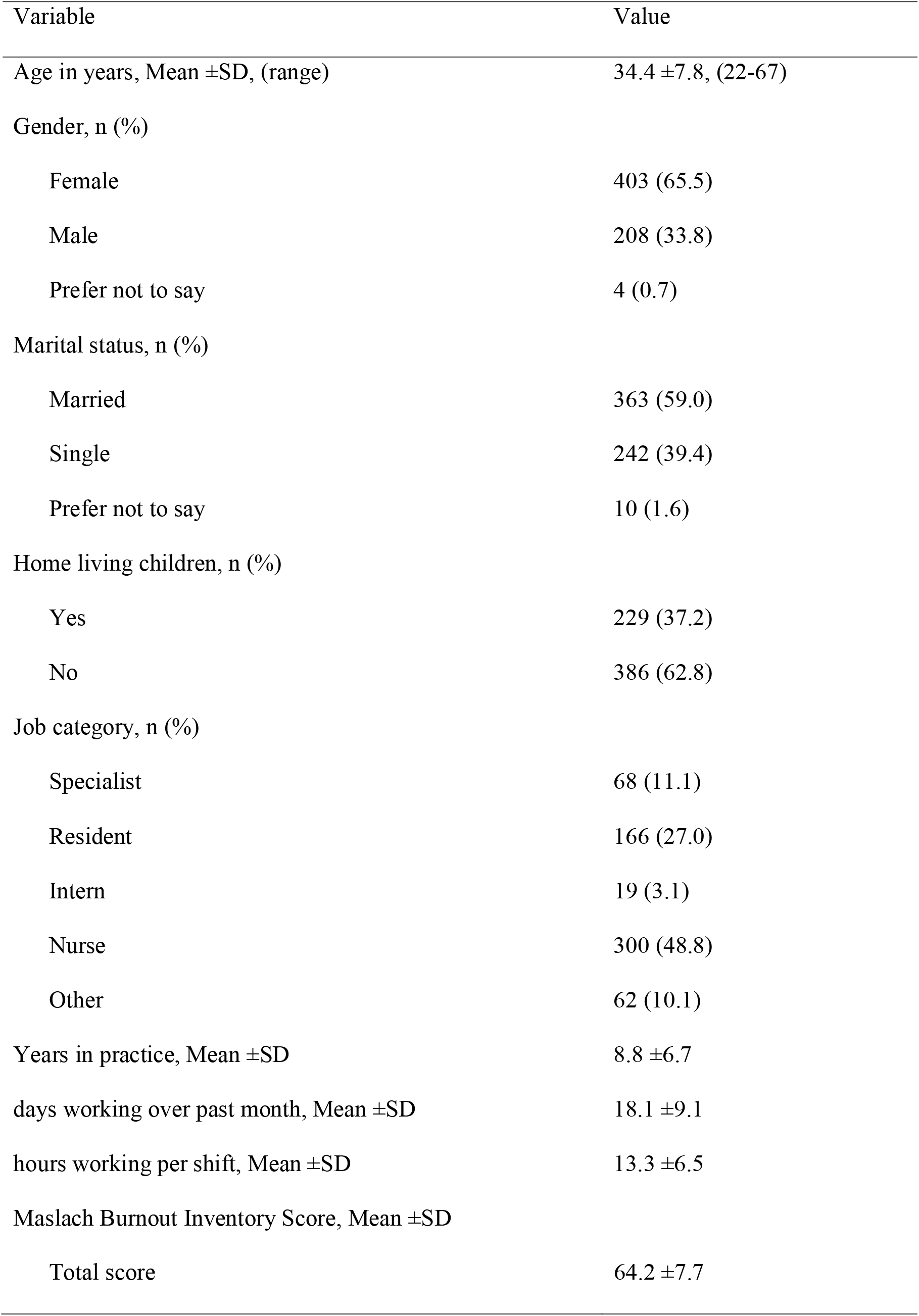
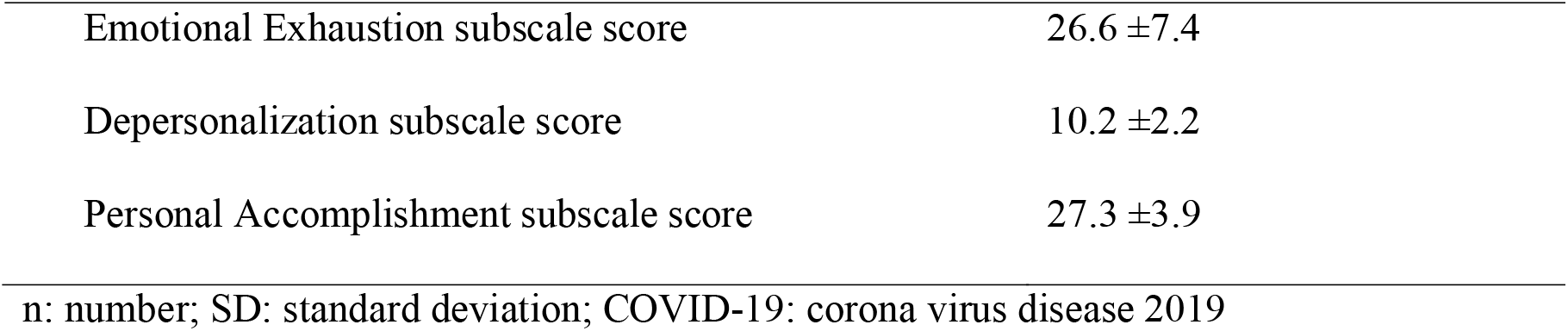
Baseline characteristics of participants (n=615)

Our findings showed that 55 (8.9%), 252 (41.0%), and 308 (50.1%) people experienced low, moderate, and high levels in the emotional exhaustion axis, respectively. In the depersonalization axis, the values for low, moderate, and high levels of burnout were 5 (0.8%), 529 (86.0%), and 81(13.2%), respectively. In addition, only one (0.8%) participant scored low in personal accomplishment axis, while 88 (14.3%) and 526 (85.5%) people were shown to experience moderate or high levels, respectively.

Table 2 illustrates how the level of burnout in each subscale varies between different groups of participants in terms of sociodemographic variables and job category. Among the studied characteristics, gender was the only variable that was associated with high levels of all three domains of burnout. High emotional exhaustion was significantly more prevalent in females and those without children, as well as people from different job categories (most common in residents and then nurses). High levels of depersonalization were significantly more common in males and younger (≤36 years) participants.

**Table 2.** Participants’ level of burnout in each dimension by sociodemographic characteristics and job category, number (%)

|  | Emotional Exhaustion |  |  |  | Depersonalization |  |  |  | Personal Accomplishment |  |  |  |
| --- | --- | --- | --- | --- | --- | --- | --- | --- | --- | --- | --- | --- |
|  | Low | Moderate | High | P | Low | Moderate | High | P | Low | Moderate | High | P |
| Age |  |  |  |  |  |  |  |  |  |  |  |  |
| ≤36 year | 36(8.9) | 154(38) | 215(53.1) | 0.105 | 3(0.7) | 338(83.5) | 64(15.8) | 0.011 | 1(0.2) | 59(14.6) | 345(85.2) | 0.723 |
| >36 year | 19(9.0) | 98(46.7) | 93(44.3) |  | 2(1.0) | 191(91.0) | 17(8.1) |  | 0 (0) | 29(13.8) | 181(86.2) |  |
| Gender |  |  |  |  |  |  |  |  |  |  |  |  |
| Female | 30(7.4) | 148(36.7) | 225(55.8) | <0.001 | 3(0.7) | 359(89.1) | 41(10.2) | 0.003 | 0(0.0) | 45(11.2) | 358(88.8) | 0.003 |
| Male | 25(12.0) | 102(49.0) | 81(38.9) |  | 2(1.0) | 166(79.8) | 40(19.2) |  | 1(0.5) | 42(20.2) | 165(79.3) |  |
| Marital status |  |  |  |  |  |  |  |  |  |  |  |  |
| Married | 40(11.0) | 142(39.1) | 181(49.9) | 0.436 | 4(1.1) | 318(87.6) | 41(11.3) | 0.077 | 1(0.3) | 57(15.7) | 305(84.0) | 0.327 |
| Single | 15(6.2) | 105(43.4) | 122(50.4) |  | 1(0.4) | 202(83.5) | 39(16.1) |  | 0(0.0) | 30(12.4) | 212(87.6) |  |
| Other | 0(0.0) | 5(50.0) | 5(50.0) |  | 0(0.0) | 9(90.0) | 1(10.0) |  | 0(0.0) | 1(10.0) | 9(90.0) |  |
| Having children |  |  |  |  |  |  |  |  |  |  |  |  |
| Yes | 24(10.5) | 104(45.4) | 101(44.1) | 0.027 | 2(0.9) | 206(90.0) | 21(9.2) | 0.033 | 1(0.4) | 43(18.8) | 185(80.8) | 0.010 |
| No | 31(8.0) | 148(38.3) | 207(53.6) |  | 3(0.8) | 323(83.7) | 60(15.5) |  | 0(0) | 45(11.7) | 341(88.3) |  |

| Job category |  |  |  |  |  |  |  |  |  |  |  |  |
| --- | --- | --- | --- | --- | --- | --- | --- | --- | --- | --- | --- | --- |
| Specialist | 7(10.3) | 31(45.6) | 30(44.1) | 0.008 | 0(0.0) | 65(95.6) | 3(4.4) | 0.657 | 0(0.0) | 8(11.8) | 60(88.2) | <0.001 |
| Resident | 7(4.2) | 59(35.5) | 100(60.2) |  | 2(1.2) | 134(80.7) | 30(18.1) |  | 0(0.0) | 7(4.2) | 159(95.8) |  |
| Intern | 1(5.3) | 14(73.7) | 4(21.1) |  | 0(0.0) | 18(94.7) | 1(5.3) |  | 0(0.0) | 3(16.8) | 16(84.2) |  |
| Nurse | 26(8.7) | 115(38.3) | 159(53.0) |  | 3(1.0) | 257(85.7) | 40(13.3) |  | 1(0.3) | 47(15.7) | 252(84.0) |  |
| Other | 14(22.6) | 33(53.2) | 15(24.2) |  | 0(0.0) | 55(88.7) | 7(11.3) |  | 0(0.0) | 23(37.1) | 39(62.9) |  |
| p: p value |  |  |  |  |  |  |  |  |  |  |  |  |

We noted that of all 615 participants in the study, who had completed the MBI questionnaire, 326 persons (53.0%) experienced high levels of burnout (defined as high levels in either domains of emotional exhaustion and depersonalization). Table 3 shows the distribution of high burnout in different groups of participants based on their sociodemographic characteristics and work-related factors.

**Table 3.**
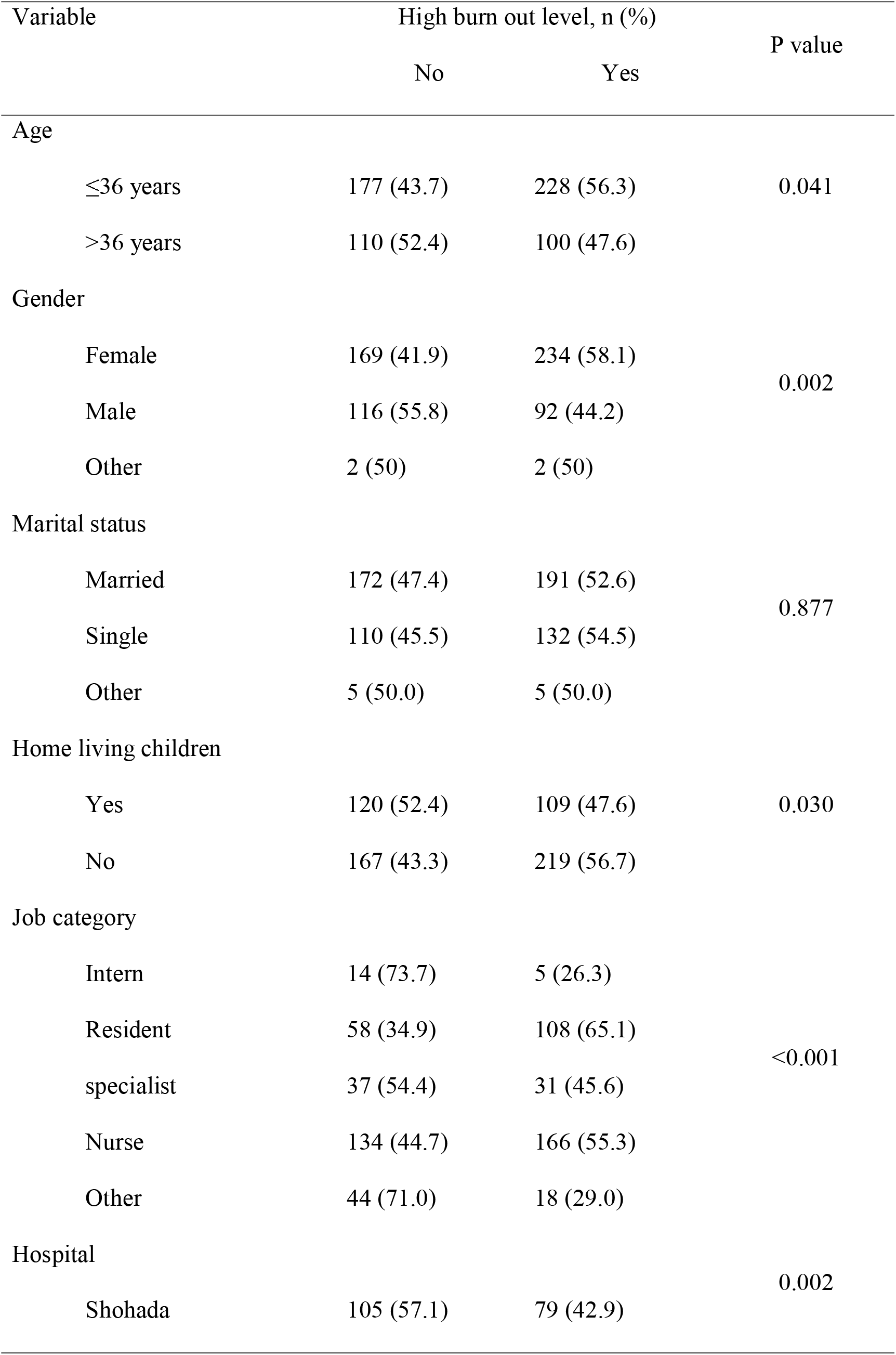
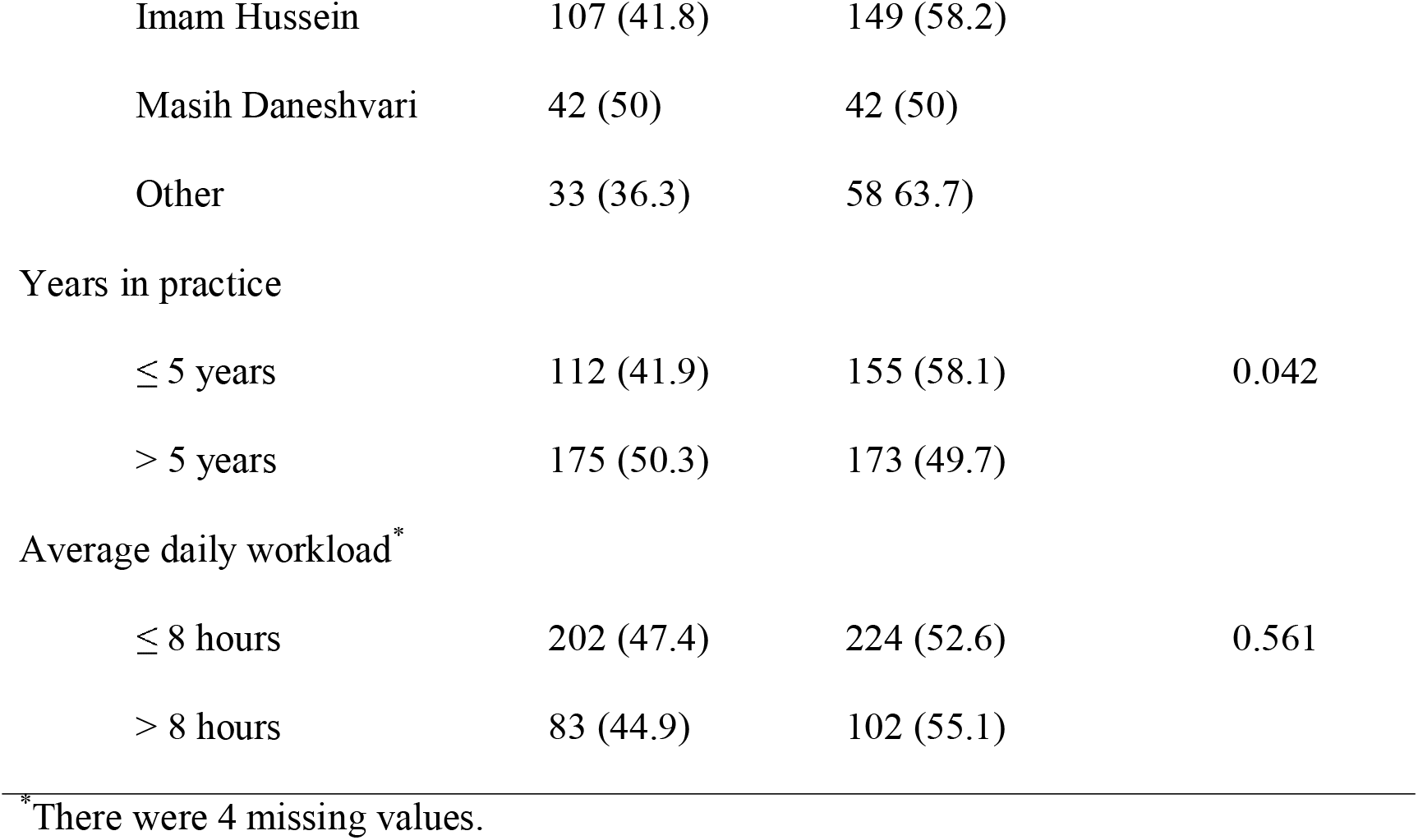
Frequency of high burnout in different groups of participants (n=615)

| Variable | High burn out level, n (%) |  | P value |
| --- | --- | --- | --- |
|  | No | Yes |  |
| Age |  |  |  |
| ≤36 years | 177 (43.7) | 228 (56.3) | 0.041 |
| >36 years | 110 (52.4) | 100 (47.6) |  |
| Gender |  |  |  |
| Female | 169 (41.9) | 234 (58.1) | 0.002 |
| Male | 116 (55.8) | 92 (44.2) |  |
| Other | 2 (50) | 2 (50) |  |
| Marital status |  |  |  |
| Married | 172 (47.4) | 191 (52.6) | 0.877 |
| Single | 110 (45.5) | 132 (54.5) |  |
| Other | 5 (50.0) | 5 (50.0) |  |
| Home living children |  |  |  |
| Yes | 120 (52.4) | 109 (47.6) | 0.030 |
| No | 167 (43.3) | 219 (56.7) |  |
| Job category |  |  |  |
| Intern | 14 (73.7) | 5 (26.3) | <0.001 |
| Resident | 58 (34.9) | 108 (65.1) |  |
| specialist | 37 (54.4) | 31 (45.6) |  |
| Nurse | 134 (44.7) | 166 (55.3) |  |
| Other | 44 (71.0) | 18 (29.0) |  |
| Hospital |  |  |  |
| Shohada | 105 (57.1) | 79 (42.9) | 0.002 |
| Imam Hussein | 107 (41.8) | 149 (58.2) |  |
| Masih Daneshvari | 42 (50) | 42 (50) |  |
| Other | 33 (36.3) | 58 63.7) |  |
| Years in practice |  |  |  |
| ≤ 5 years | 112 (41.9) | 155 (58.1) | 0.042 |
| > 5 years | 175 (50.3) | 173 (49.7) |  |
| Average daily workload* |  |  |  |
| ≤ 8 hours | 202 (47.4) | 224 (52.6) | 0.561 |
| > 8 hours | 83 (44.9) | 102 (55.1) |  |

\*There were 4 missing values.

The results of multiple logistic regression analysis of different variables to detect their impact on the development of high burnout levels have been presented in Table 4. As can be seen in the table, of different variables included in analysis, age, gender, job category and hospital were significantly associated with burnout status; while marital status, having children, number of years in practice, and the average daily workload were not associated with this outcome.

**Table 4.** Bivariate associations between high level of burnout and sociodemographic and work-related factors (n=611)

| Variable | OR | 95% CI for OR |  | P value |
| --- | --- | --- | --- | --- |
|  |  | Lower limit | Upper limit |  |
| Age<sup>*</sup> | 0.97 | 0.95 | 0.99 | 0.010 |
| Gender |  |  |  |  |
| • Female | 1 |  |  | 0.015 |
| • Male | 0.65 | 0.45 | 0.92 |  |
| Job category |  |  |  | <0.001 |
| • Intern | 1 |  |  |  |
| • Resident | 6.64 | 2.16 | 20.41 | 0.001 |
| • specialist | 4.13 | 1.18 | 4.40 | 0.026 |
| • Nurse | 4.56 | 1.54 | 13.48 | 0.006 |
| • Other | 1.77 | 0.53 | 5.90 | 0.350 |
| Hospital |  |  |  | 0.011 |
| • Shohada | 1 |  |  |  |
| • Imam Hussein | 1.86 | 1.25 | 2.78 | 0.002 |
| • Masih Daneshvari | 1.29 | 0.74 | 2.23 | 0.364 |
| • Other | 1.96 | 1.08 | 3.53 | 0.026 |

n: number; yr: year; SD: standard deviation; CI: confidence interval; OR: odds ratio
<sup>\*</sup>Per 1 year increase

## Discussion

The results of this cross-sectional survey demonstrated high levels of burnout in healthcare workers looking after COVID-19 patients. We also found that younger age and female gender were predisposing factors for burnout and that the level of burnout varied significantly by site of practice and job category (with residents being the most vulnerable group).

The results of this study showed that high levels of burnout are present among healthcare professionals who take care of COVID-19 patients. Other studies have also investigated the psychological impact of COVID-19 and other infectious disease outbreaks on hospital staff. A recent rapid review of the existing literature retrieved 59 papers that described the emotional reactions of healthcare providers dealing with patients during an outbreak of viral infectious disease. The investigators concluded that staff in contact with these patients had greater levels of acute or posttraumatic stress (odds ratio 1.71, 95% confidence interval 1.28 to 2.29) and psychological distress (1.74, 1.50 to 2.03) as compared with lower risk controls (17). Most of these studies used instruments other than MBI, and only eight were on COVID-19. Poon et al, using State-Trait Anxiety Inventory, conducted a survey to identify anxiety levels among 1926 front-line health care workers in Hong Kong during SARS outbreak. They also used the emotional exhaustion dimension of the MBI. Mean anxiety levels (51.1 versus 47.1; P<0.001) and the number of burnout symptoms (7.3 versus 5.1; P<0.001) among front-line health care workers were higher than those among controls; anxiety scores correlated with burnout scores (18). In another study by Fiksenbaum and colleagues, an on-line survey of 333 nurses (315 women, 18 men) working in the province of Ontario, Canada was performed to investigate the role of perceived SARS threat and organizational support in predicting emotional exhaustion and stated anger (19). In their study, mean emotional exhaustion score was 4 (±1.85). In a cross-sectional survey conducted in China, using General Health Questionnaire, Dai et al evaluated mental health status of a convenient sample of 4357 healthcare workers (20). They found that 1,704 workers (39.1%) had psychological distress (score≥3). In another survey, Lai and colleagues assessed the magnitude of mental health outcomes of 1257 healthcare workers treating patients exposed to COVID-19 in 34 hospitals in China. They found that 634 people (50.4%) reported symptoms of depression, 560 (44.6%) anxiety, 427 (34.0%) insomnia, and 899 (71.5%) distress (21). Moreover, Liu and colleagues, using the Zung Self-rating Anxiety Scale, estimated the prevalence of anxiety among medical staff to be 12.5% (22). Xing et al in a cross-sectional survey using Symptom Checklist-90 (SCL-90) investigated the mental health status of 548 medical personnel dealing with COVID-19 at 12 hospitals in eight provinces and cities of China (23). According to the findings of theyr study, overall average score of SCL-90 and mean values of factors (somatization, obsessive-compulsive, anxiety, phobic anxiety, and psychoticism) among medical personnel were significantly higher than that of the norm group. Zhu et al evaluated stress, depression, and anxiety among 5062 health workers using Impact of Event Scale-Revised, Patient Health Questionnaire-9, and Generalized Anxiety Disorder 7-item. In their study 29.8 percent of participants reported stress, 13.5% depression, and 24.1% anxiety (24). In a study by Maunder on 587 workers at Toronto Hospital, 30.4% had high emotional exhaustion (score≥27) (25). To the best of our knowledge, no prior study used MBI to assess the psychological sequel of COVID-19; therefore, a comparison of the results of this study with that of previous studies is not feasible. Burnout is a very well known consequence of healthcare for the staff and is strongly linked to significant outcomes such as patient safety (11,12); Moreover, MBI is a validated instrument to assess the level of burnout. Therefore, we believe that MBI is a practical and credible method for assessment of the psychological impact of COVID-19 pandemic on staff.

Our study lacked a control group but a comparison of our findings with other studies on burnout in our country before the COVID-19 pandemic shows a much higher level of burnout. A systematic review of the studies which examined the prevalence of burnout among nurses in Iran, published in 2018, found that the overall prevalence of burnout among Iranian nurses was 36% (95% CI, 20-53%) (26). In our study, more than 55% of the nurses experienced high burnout during the COVID-19 pandemic. In a cross-sectional study conducted on 208 primary health care providers in Iran, the average score of the participants in emotional exhaustion dimension was 17.19, while in our study the average score was around 27 in this subscale (27). A large study to assess the level of job burnout among 1807 healthcare providers in Iran found that mean scores (±SD) in emotional exhaustion, depersonalization, and personal accomplishment subscales were 8.9 (± 9.0), 23 (± 2.9), and 34 (± 8.6), respectively (28); while in our study these values were much higher: 26.6 (±7.4), 10.2 (±2.2), and 27.3 (±3.9), respectively.

Several outbreaks of infectious diseases have occurred over the past two decades and they represent a serious threat to both health services and staff. Healthcare providers, who need to take care of a large number of potentially infectious victims, are under great physical and psychological pressure. Therefore, it is not surprising to expect a high rate of infection as well as mental health problems among the staff. Chen et al interviewed 13 medical staff at a hospital in China during the COVID-19 outbreak and noted that “getting infected was not an immediate worry to staff” but that “they were afraid of bringing the virus to their home”. They also stated that issues such as ambiguity regarding “how to deal with patients when they were unwilling to be quarantined at the hospital or did not cooperate with medical measures because of panic or a lack of knowledge about the disease” and “shortage of protective equipment” were also contributing to poor mental health of medical staff (29). While our study was not meant to discover the reasons for this phenomenon, several issues can be speculated to have resulted in this state of high burnout. Heavy workload, fear of contracting the disease or transmitting it to the family, as well as lack of staff support systems are often cited complaints. Whether these are linked to the development of high burnout or not and the contribution of each factor can be the subject of future studies. This will help administrators adapt the best approach to maintain staff mental health.

Our study found age and gender to be important predisposing factors for high burnout. Several studies have shown that staff who are women (3,21,24,30-32) or younger (33-37) are more vulnerable to psychological distress. Our findings are in accordance with these studies. However, in two studies on personnel taking care of patients with covid-19, older age was a risk factor for psychological symptoms (23,36).

While some studies have found out that having children (38,39) and having an increased contact with affected patients (18-21,25,30-32,34-36,38-45,47,48) are predisposing factors, our study failed to find such a relationship. Moreover, several studies have shown that being less experienced (3,30,42,47) puts the staff at higher risk. While we found a significant difference between the experience of those with and without high burnout, this variable did not remain in the final regression model, pointing to potential co-linearity between age and experience.

Within the job categories, residents were found in this study to be at higher risk of developing high burnout. Nurses were the second at-risk group. In previous studies, nurses were generally at higher risk than doctors (18,21,34,35,38,39,48,49). Lai et al, for example, reported more severe degrees of all measurements of mental health symptoms in nurses exposed to COVID-19 (21). However, two studies reported the opposite (50,51). A survey of 99 residents during AH1N1 influenza virus outbreak in a third level hospital in Mexico City conducted in 2009 documented high levels of burnout among this population (33). The reasons why residents were so vulnerable to burnout needs further study, but issues such as heavy workload, changes in duty rosters to accommodate the new needs, and cancellation of vacations, as well as having less access to personal protective equipment may have caused this unfavorable state.

The level of burnout in our study was also affected by the location of work. In a study by Maunder on 769 healthcare workers during SARS outbreak, the investigators found a significantly higher levels of burnout in the staff of one of the two studied hospitals (median score 19, interquartile (IQR) range 10–29, in one hospital versus median score 16, IQR 9– 23, in eh other hospital) (25). Dai et al found that working in primary hospitals were poor prognostic factors (20). In our study, working in the hospitals which were not designated as COVID-19 centers was accompanied by higher levels of burnout. Several reasons might account for this difference: working conditions affects perceived threat; designated hospitals are better prepared and equipped for healthcare delivery to COVID-19 patients; working in non-designated hospitals is not rewarded by recognition and incentives found in COVID-19 designated hospitals.

### Strengths and limitations

Our study has several strengths. Burnout is a well recognized psychological consequence of working in the healthcare section and its impact on staff physical and emotional health, patient safety, and quality of care is documented. The instrument used in this study is well validated across geographical areas. We used a relatively large sample with an acceptable response rate.

This study also faces some limitations. Although we tried to consider several personal (e.g. marital status and having dependent children) and work-related parameters (e.g. workload, job category), our study might have failed to deal with all confounding factors. We specifically referred to COVID-19 in our questions; however, a comparison with other clinicians or the general population has not been performed and we cannot claim that this level of burnout is solely attributable to COVID-19 pandemic.

### Suggestions for future research

Other studies, using qualitative methods, are necessary to delve into the reasons for this observed phenomenon. In-depth interviews can bring important issues into consideration, which can be the subject of further studies. The necessity of implementation and evaluation of various measures and strategies to overcome this high prevalence of burnout among staff, through interventional studies, cannot be overemphasized. Last but not least, further assessment of the long term sequel of COVID-19 pandemic through follow up studies deserves particular attention.

### Conclusion

Our study highlighted the high prevalence of burnout among healthcare workers who are caring for patients during the new pandemic. Several personal and work related factors contribute to the level of burnout that the staff experience. These include younger age, female gender, working as a resident or nurse, and working in non-COVID-designated hospitals.

## Data Availability

Data available.

